# Predicting onward care needs at admission to reduce discharge delay using machine learning

**DOI:** 10.1101/2024.08.07.24311596

**Authors:** Chris Duckworth, Dan Burns, Carlos Lamas Fernandez, Mark Wright, Rachael Leyland, Matthew Stammers, Michael George, Michael Boniface

## Abstract

Early identification of patients who require onward referral for social care can prevent delays to discharge from hospital. We introduce a machine learning (ML) model to identify potential social care needs at the first point of admission. The model performance is comparable to clinician’s predictions of discharge care needs, despite working with only a subset of the information available to the clinician. We find that ML and clinician perform better for identifying different types of care needs, highlighting the added value of a potential system supporting decision making. We also demonstrate the ability for ML to provide automated initial discharge need assessments, in the instance where initial clinical assessment is delayed. Finally, we demonstrate that combining clinician and machine predictions, in a hybrid model, provides even more accurate early predictions of onward social care requirements and demonstrates the potential for human-in-the-loop decision support systems in clinical practice.

## Introduction

A critical task for hospital care teams is to promptly identify patients who require onward care support. For patients in need of onward care, whether at home or in care homes, any delays in assessment, choice, or access prevent discharge from hospital^1^. Delays to discharge lead to poor patient outcomes, including increased risk of hospital-acquired infection, decreased mobility, physical deterioration, worsening mental health, and cognitive deconditioning^2-5^. Operationally, medically fit patients occupying bedspace create substantial problems for patient flow; increasing pressure across ambulatory services and urgent care systems^6^. In the UK, the National Health Service (NHS) is under significant pressure, with a backlog of 7.6 million patients waiting for treatment and a median waiting time of 15 weeks^7^. Bedspace is at critical levels, with occupancy consistently exceeding 93% through last winter in NHS England^8^. As of January 2024, 14,436 patients a day (14% of bed capacity) remained in hospital while being ready to leave^9^.

Despite the majority of patients being discharged without the need for additional support (85% as of December 2022^6^), the driving reason for discharge delay is the need for onward care following an illness or injury. For patients in UK hospitals for seven days or more, 65% of discharge delays are attributed to waiting for arrangement of care at home (25%), short-term reablement (22%) and permanent care or a nursing home (18%). In NHS England, these onward needs are organised into four distinct ‘Discharge Pathways’ as shown in Figure 1. Community services face significant issues with both staffing and funding, ultimately leading to problems with capacity and response times^1,10^. Internal processes at hospitals also cause delay, including delay in assessment or decision-making, misidentification of care requirements, transfer between clinical specialities^11^, and lower discharge rates at weekends^12^. It is critical that care requirements can be assessed and identified as early as possible to avoid delay. In-line with University Hospitals Southampton (UHS) NHS standard operating procedure, an initial discharge assessment should be made within the first 24 hours of hospital admission^13^.In practice, this initial planning is provided for less than 50% of patients. Awaiting completion of assessment was listed as the primary reason for discharge delay for 20% of NHS patients^14^, likely exacerbated by workload and inability to identify potential care requirements.

**Figure 1:**
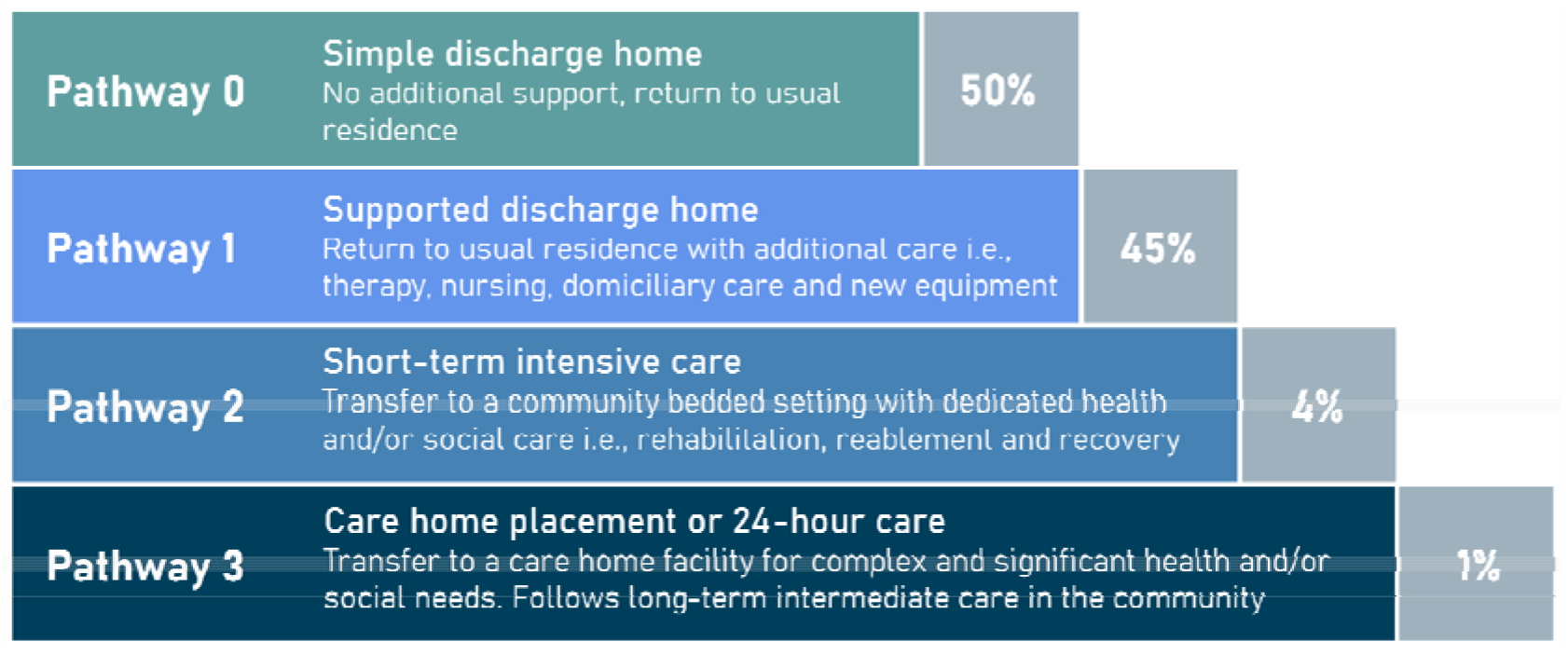
Discharge pathways of increasing health and social care requirements. Percentages represent NHS targets for patients discharged on each pathway.

The aim of this paper is to demonstrate the potential for machine learning to support the early and accurate identification of onward care needs. Early identification creates the possibility to release a large proportion of capacity back into the system (both care hours wasted when a patient is too unwell to leave the hospital, and in social worker hours collating details regarding the patient’s most current functional ability).

First, we introduce an explainable machine learning model to predict patients’ potential discharge care needs at the point of admission to hospital. Learning from historical best practice, supervised machine learning has been validated in a variety of clinical settings^15^ including cancer diagnosis^16^, sepsis prediction^17^ and emergency admissions^18^. The prediction is made based on basic, digitally available patient information (e.g., age, arrival mode, admission route, chief complaint, history of prior spells and prior discharge needs) at the first point of admission. Using machine learning explanations, we can also identify the most important factors to review when assessing a patient’s onward care needs.

Second, we consider a human-machine hybrid model which updates the machine learning prediction based on the clinician’s own initial assessment of discharge requirements. Machine learning algorithms naturally have capacity to capture more complex trends in data but are intrinsically limited by the inability to capture important contextual domain knowledge (e.g., visual cues of patient or conversation with patient or family). A hybrid approach combining human and machine predictions integrates both contextual domain knowledge and complex data mining to best identify potential discharge requirements and reduce likelihood of delay.

## Methods

### Data

We used a pseudonymised version of routinely collected data on patient admissions and hospital spells at UHS between 1^st^ January 2017 and 1^st^ January 2023. Cohort selection is described in Figure 2, which we briefly summarise here. Patient history is computed from 757,421 spells (including 503,542 outside of study period). Discharge planning information is managed by the Application Express (APEX) system which was introduced into UHS working practice as of 19^th^ August 2019 before being widely adopted at the Trust by 1^st^ June 2020. APEX records discharge planning information, including expected discharge date, predicted discharge pathway and additional notes about complex care needs. This accounted for 94,490 spells from which patient discharge history is computed. The main cohort used in modelling is 51,754 once short spells and those without planning information are removed.

**Figure 2:**
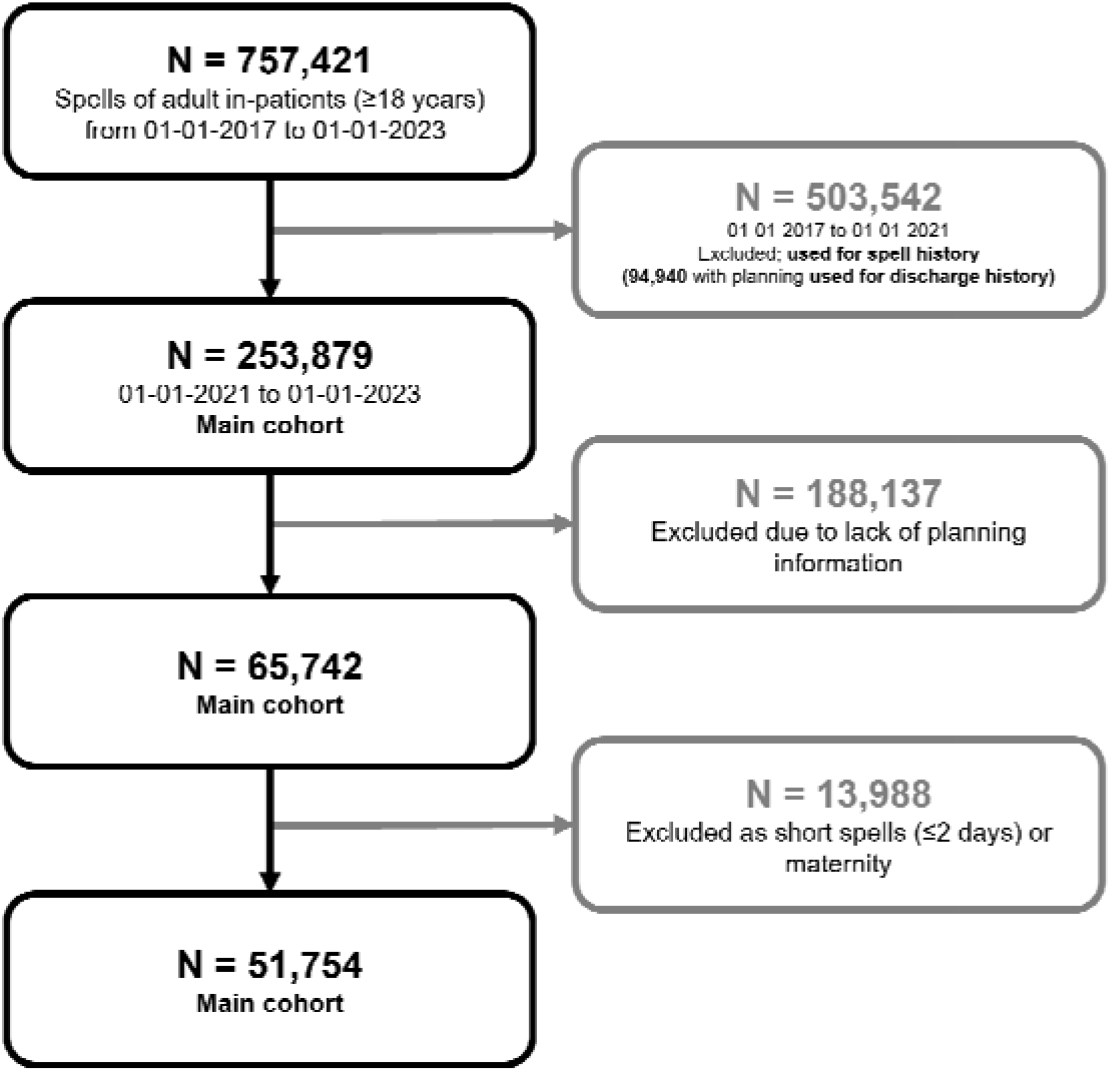
Cohort selection diagram. Darker arrows and boxes correspond to selection of the core cohort, while lighter arrows and boxes (grey) correspond to excluded spells (which may be used for auxiliary purposes, as described).

We use the NHS England Discharge to Assess pathways as a multi-class target for our machine learning models (as shown in Figure 1). Introduced as best practice in 2016, this framework ranges from reablement and rehabilitation to short-term care, with longer-term needs for care and support assessed at a point of optimal recovery^19,20^. Other than in exceptional circumstances, no patient should be discharged directly into a permanent care home for the first time without first exhausting home first as a least restrictive discharge venue or discharging into a temporary placement. These pathways represent an ordinal mapping from care acuity and are encoded 0-3 when used in feature engineering to summarise patient’s discharge histories. We take the last discharge pathway registered in APEX by the clinical team before discharge. In Figure 2, we show the actual distribution of discharge pathways throughout the study period for training and validating our machine learning model. The overall distribution of pathways is significantly different to NHS guidelines as found in Figure 1 (i.e., Pathway 0: 69.4%, Pathway 1: 12.8%, Pathway 2: 10.9%, Pathway 3: 7.0%). In the right panel of Figure 2, we note a shift in the proportion of spells discharged on Pathways 1 and 3 around March 2022, likely reflective of discharge policy change.

For our study cohort (i.e., period in which we train and validate our models), we select spells that occurred between 1st January 2021 and 1st January 2023, with corresponding APEX discharge planning information. This ensures we can collect patient history over a 4-year baseline and a discharge history over the prior six months. We remove short spells (i.e., less than two days) leaving 51,754 for modelling. Most short spells do not require discharge planning and hence are not the target for prediction. We note, however, that including spells (≤2 days) does not significantly change the performance of our model. The general characteristics of our study cohort stratified by discharge pathway are given in Table 1.

**Table 1:** Descriptive statistics of the study cohort spells stratified by final discharge pathway. Values are spell count (percentage). Percentage is normalised by column-wise (i.e., by discharge pathway).

|  | Pathway 0 | Pathway 1 | Pathway 2 | Pathway 3 |
| --- | --- | --- | --- | --- |
| **Age Group** |  |  |  |  |
| 18-44 | 6333 (17.7%) | 209 (3.17%) | 99 (1.76%) | 181 (5.02%) |
| 45-64 | 11385 (31.9%) | 929 (14.1%) | 558 (9.92%) | 406 (1.13%) |
| 65-84 | 15955 (44.6%) | 3302 (50.0%) | 3039 (54.0%) | 1638 (45.4%) |
| 85+ | 2062 (5.77%) | 2162 (32.7%) | 1928 (34.3%) | 1383 (38.3%) |
| **Gender** |  |  |  |  |
| Female | 16260 (45.3%) | 3751 (56.8%) | 3212 (57.1%) | 2029 (56.1%) |
| Male | 19644 (54.7%) | 2855 (43.2%) | 2414 (42.9%) | 1585 (43.8%) |
| **Elective Spell** |  |  |  |  |
| Yes | 9059 (25.2%) | 251 (3.80%) | 279 (4.96%) | 78 (2.16%) |
| No | 26848 (74.8%) | 6355 (96.2%) | 5347 (95.0%) | 3537 (97.8%) |
| **Admission via ED** |  |  |  |  |
| Yes | 17745 (49.4%) | 5450 (82.5%) | 4493 (79.9%) | 3019 (83.5%) |
| No | 18162 (50.6%) | 1156 (17.5%) | 1133 (20.1%) | 596 (16.4%) |
| **Has Spell History** |  |  |  |  |
| Yes | 23433 (65.2%) | 5861 (88.7%) | 4454 (79.2%) | 2951 (81.6%) |
| No | 12474 (34.7%) | 745 (11.2%) | 1172 (20.8%) | 664 (18.3%) |

| Has Discharge History |  |  |  |  |
| --- | --- | --- | --- | --- |
| Yes | 7863 (21.9%) | 3562 (53.9%) | 2183 (38.8%) | 1706 (47.2%) |
| No | 28044 (78.1%) | 3044 (46.0%) | 3443 (61.1%) | 1909 (52.8%) |

Predictions of discharge pathways are made at the very first point of admission (i.e., before information relating to care speciality or planned treatment is made available), with each patient characterised by basic patient descriptors (e.g., age, ethnicity), admission method (e.g., if the patient was admitted as GP referral or through ED), information collected in the emergency department (if applicable) and histories from prior spells and discharge plans. Prediction at this stage helps early identification of elevated onward care needs, ensuring transfer of care teams can triage earlier and have additional time to organise social care, which has significant lead times.

All numerical variables were used directly as input to the model, whereas categorical variables are target-encoded^21,22^. We provide a more detailed description of our encoding in Supplementary Methods Section 1.1 and the full list of features in Supplementary Table 1. In Figure 3 we show the distribution of the most prevalent categories for a selection of features used in modelling, stratified by the final discharge pathway for the spell. The electronic health records used by our models are available to review by clinicians and are used in regular practice. Our model did not have access to any free text fields in the electronic health records. Analysis was performed in Python 3.9.5 using standard packages including numpy^23^, matplotlib^24^ and pandas^25^.

**Figure 3:**
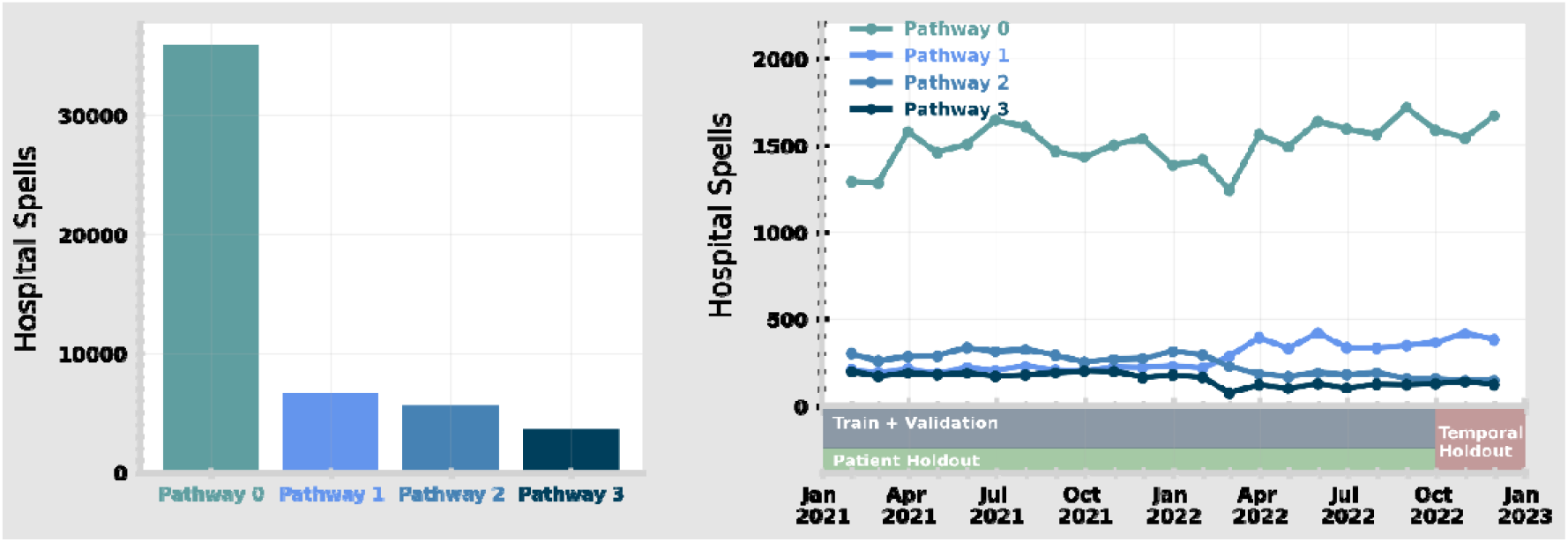
Overall distribution of discharge pathways (left) and monthly average for each pathway (right) throughout the study period. Visual representations of the separation of the study data into training, patient test and temporal test sets is also given on the right.

**Figure 4:**
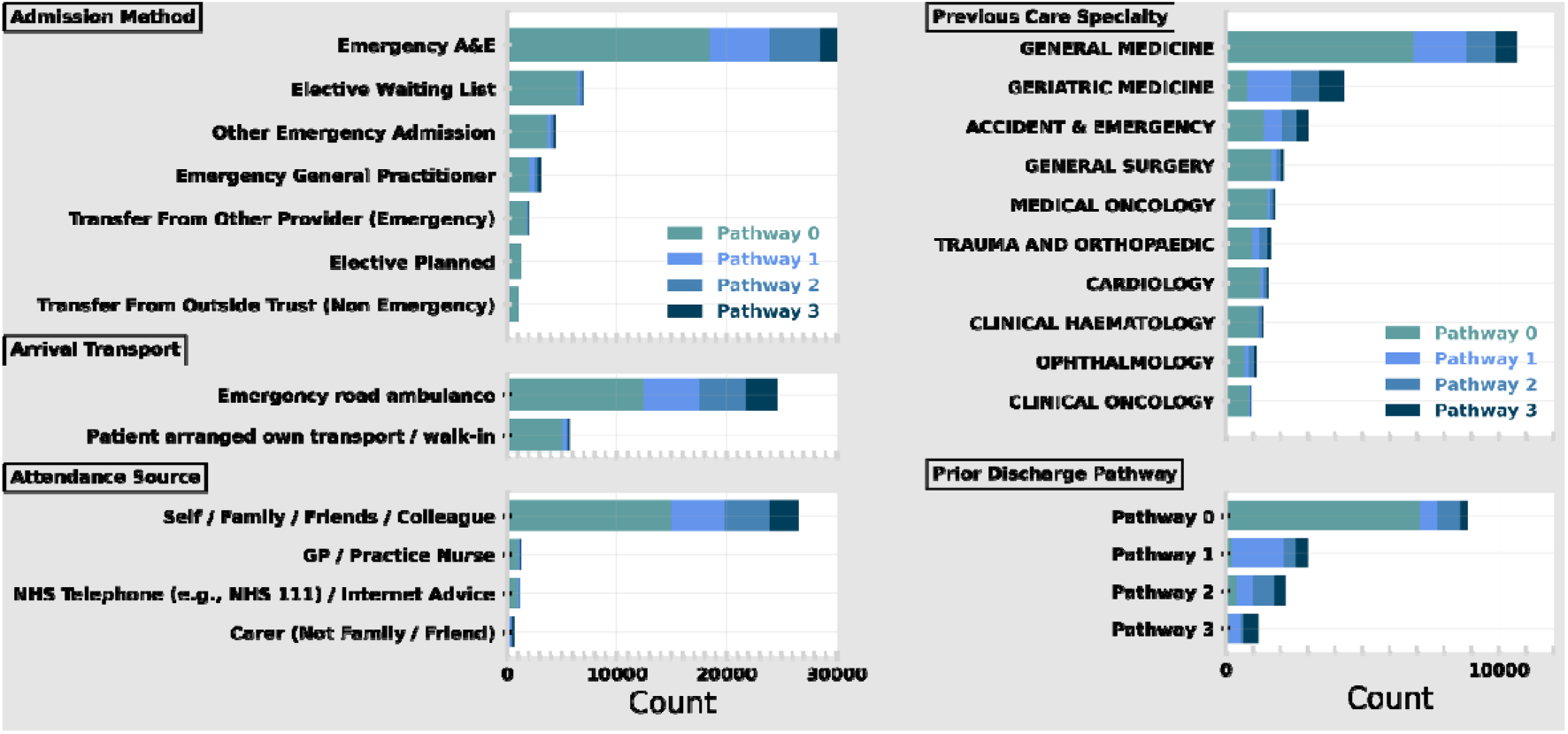
Frequency of most common categories in the following features: Admission Method (top left), Arrival Transport (middle left), Attendance Source (bottom left), Previous Care Speciality – i.e., from prior spell (top right) and Prior Discharge Pathway (bottom right). The stacked bar shows the total frequency of the category with each bar stratified by final pathway.

### Ethics and data governance

This study was approved by the University of Southampton’s Ethics and Research Governance Committee (ERGO, and approval was obtained from the Health Research Authority. All methods were carried out following relevant guidelines and regulations associated with the Health Research Authority, and NHS Digital. The research was limited to the use of previously collected, non-identifiable information. Informed consent was waived by the University of Southampton’s Ethics and Research governance committee, University of Southampton, University Road, Southampton, SO17 1BJ, United Kingdom and the Health Research Authority, 2 Redman Place, Stratford, London, E20 1JQ, UK. Data was pseudonymised (and, where appropriate, linked) before being passed to the research team. The research team did not have access to the pseudonymisation key.

### Modelling

We trained our classifier based on 29,084 unique spells (56.2% of total) and defined a validation set (7272; 14.0%) grouped at the patient level. To quantify the performance of our classifier we define two holdout test sets to consider how the model would operate in different circumstances. First, we define a patient holdout test set (i.e., multiple spells relating to a patient cannot be split between holdout and training). This contains 9177 spells (17.7%) from 1^st^ January 2021 to 30^th^ September 2022. Second, we define a 3-month temporal holdout (6221; 12.0%) which contains all spells from 1^st^ October 2022 to 1^st^ January 2023. This is designed to test the real-world performance of the model (i.e., where it is trained on historical hospital data and deployed in the same environment). While the temporal holdout best emulates how a model may be deployed, there is a risk of data leakage (i.e., if multiple spells of a given patient are correlated). A patient holdout tests the performance for patients whose prior spells were not included in the original training. For readability, we follow the results for our patient holdout test set in the main text, while also presenting results of the temporal holdout test set in the Supplementary Materials.

For our machine learning model, we used XGBoost with a ‘multi-softmax’ objective^26,27^.Gradient-boosted trees are a popular choice for large tabular data, which includes a variety of continuous, binary, and categorical data. Tree-based models can also naturally deal with missing data consistently (i.e., through the creation of splits of whether data is missing or not rather than imputation), which is important for routinely collected clinical data, as here. Despite gradient-boosting algorithms excelling at extracting signal from data, they are prone to overfitting. To help avoid this, model hyperparameters were selected using five-fold cross-validation grouped at the patient level (i.e., individual patients cannot appear in different folds). On training, we also defined 50 early stopping rounds (i.e., when the train set performance metrics become better than the validation set) to prevent overfitting. We provide more detailed information on our choice of XGBoost and hyperparameter tuning^28^ in Supplementary Materials Section 1.2 with Supplementary Table 2 giving the final hyperparameters.

Model performance was evaluated independently for both temporal and patient holdout test sets using Micro F1 Score and One-Vs-Rest (OVR) Area Under the Receiver Operating Curve (AUROC) and Average Precision (AP) for each target class.

### Model Explanations

An important part of machine learning in a clinical setting is the ability to attribute why a given recommendation has been made to the user. Explanations enable users to understand the machine learning decision making process and helps identify data which should be reviewed before making the final decision. While machine learning models are often described as ‘black boxes’, there are various methods to interpret and explain the predictive process.

We attribute the fractional contribution of each input feature to individual pathway predictions using Shapley values (i.e., Shapley Additive exPlanations; SHAP^29,30^). One strength of SHAP is that explanations are 1) locally accurate (i.e., each pathway prediction can be explained) and 2) globally consistent (i.e., explanations can be consistently aggregated across multiple predictions). This flexibility is critical in a medical environment where explanations are important on a local (i.e., for each patient) and global (i.e., to understand the operating circumstances of the model) level. We use the TreeExplainer^31,32^ implementation of SHAP designed for tree-based algorithms.

### Clinical Pathway Updates

As part of the standard operating procedure at UHS, an initial discharge assessment should be made within the first 24 hours of hospital admission, usually as part of the first consultant review^13^. In order to compare how the proposed machine learning model performs in relation to current practice, we consider all pathway assessments made within the first 24 hours post-admission by clinical care teams. In practice, this corresponds to all pathway updates registered on the APEX system. Multiple pathway updates within 30 minutes of each other are combined (retaining the most recent) as they are more likely to reflect administrative updates (e.g., opening the patient record with incomplete information) rather than distinct assessments. For the patient holdout test set this corresponds to 4283 updates across 4076 spells (46.67%). For the temporal holdout test set (Results in Supplementary Materials) this corresponds to 2845 total updates for 2757 spells (45.73% of test set).

### Hybrid Model

Machine learning is limited relative to humans by not being able to capture important contextual domain knowledge that is not easily captured in data (e.g., visual cues of a patient’s current state, conversations with family or carers). Despite this, machine learning models capture more complex trends in data than humans can retain, offering consistency and nuance in predictions. A hybrid approach combining human and machine predictions integrates domain knowledge and complex data mining to deliver a more powerful recommendation.

For spells with an initial pathway prediction, a ‘hybrid’ recommendation based on this input and the machine learning class probabilities can be made. For this model, we made use of the Scikit-Learn implementation of a Logistic Regression, adapted to make multi-class predictions^22^. Logistic regression is a linear model which aims to classify outcomes through probabilities estimated by the logistic function. We implement optimised for ≤100 iterations using an L2 penalty with class weights inversely proportional to class frequencies. For both the XGBoost and ‘hybrid’ logistic regression models we define class weights inversely proportional to the number of the samples in a target class. This ensures our classifiers are tuned to be robust against class imbalance and are calibrated to better identify higher acuity pathways.

## Results

### Machine learning model performance

In Figure 5, we consider the performance of our XGBoost model stratified by each target class. For each pathway, performance is evaluated using the Receiver Operating Curve (ROC; left) and the Precision-Recall (PR; right) Curve using a One-Vs-Rest strategy (i.e., each class is considered as binary with the rest of the predictions of the negative outcome). For both patient (top) and temporal (bottom) holdout test sets, performance is generally better for lower acuity care pathways (i.e., no care needs easier to identify than elevated care needs).

**Figure 5:**
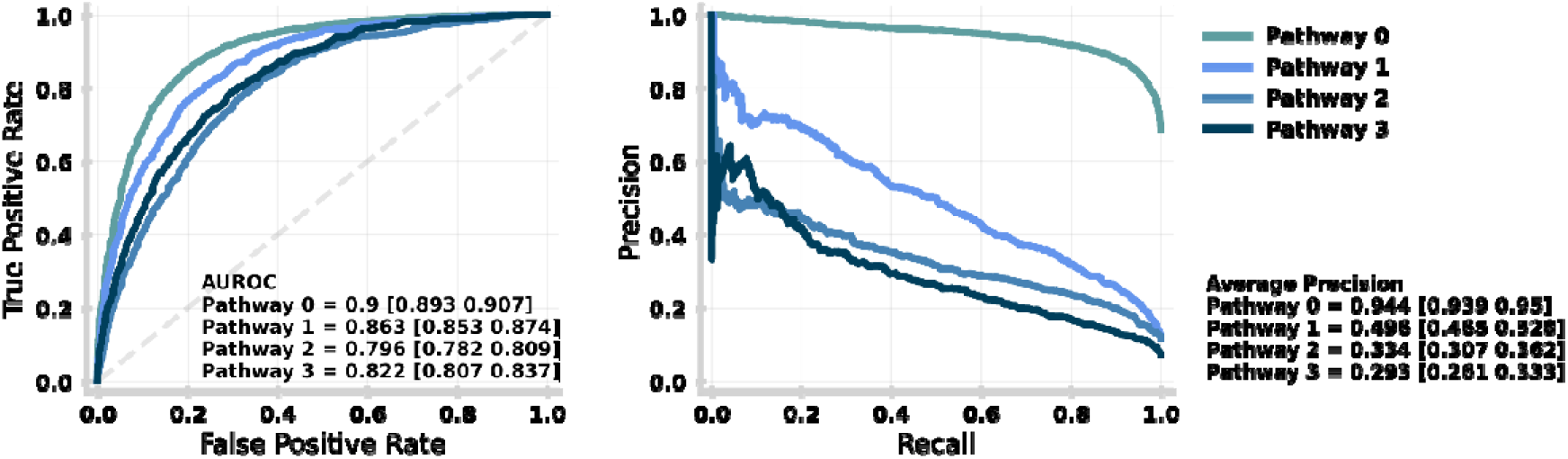
Performance of the machine learning model stratified by target class (i.e., discharge pathway) for the patient holdout test set. The performance for each target class is given by the receiving operator curve (left) and precision-recall curve (right) with the Area Under the ROC and Average Precision (over all recall thresholds) annotated respectively with 95% confidence intervals given in square brackets. Both ROC and PR are evaluated using a One-vs-Rest (OVR) strategy for each target class. An equivalent plot for the temporal holdout test set is given in Supplementary Figure 2.

Performance stratified by pathway is shown further in Figure 6, which contains the confusion matrix for the patient holdout test set. Despite implementing sample weights (to balance the importance of predictions across pathways), the XGBoost model is best at identifying patients to be discharged on Pathway 0. Regardless, the model performs well at identifying patients in need of needing any elevated care needs (i.e., predicting pathways 1-3) as denoted by the PR curve of Pathway 0 in Figure 5.

**Figure 6:**
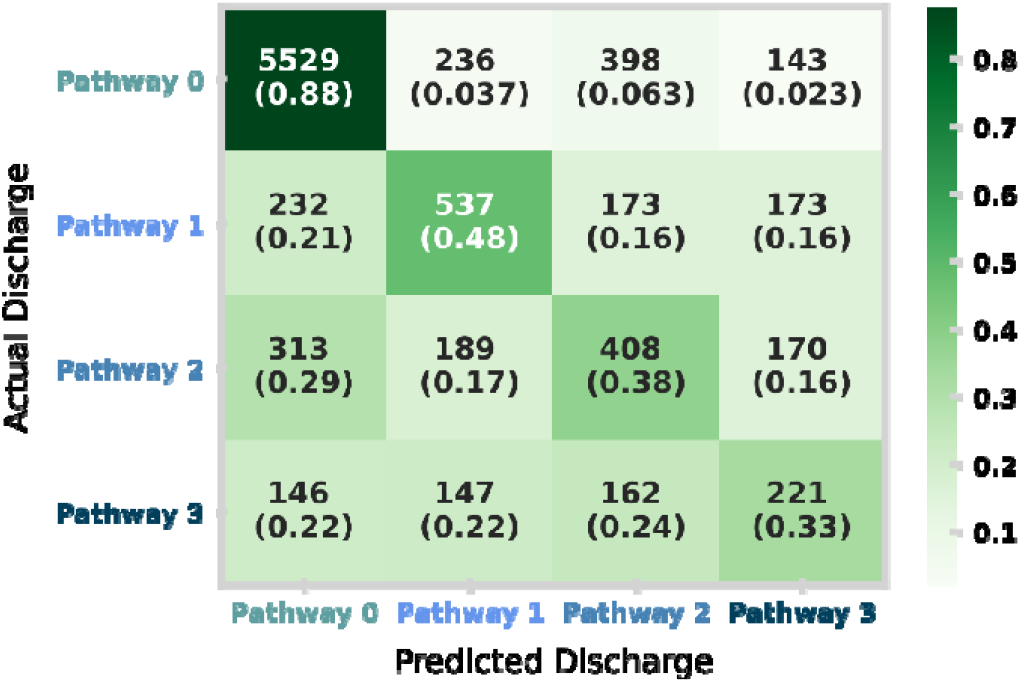
Confusion matrix for the patient holdout test set. The matrix shows the distribution of model prediction (columns) against the actual discharge (rows). In each square, the total counts and row normalised (i.e., by actual discharge) proportions are shown. An equivalent plot for the temporal holdout test set is given in Supplementary Figure 3.

In Figure 6, we show the confusion matrix for the patient holdout test set demonstrating our model to be well tuned to identify higher acuity pathways. Despite this, we note distinct differences in class mixing between the patient holdout and the temporal holdout (Supplementary Figure 2). We find the temporal holdout has a slight bias towards Pathway 1 with potential data drift towards the end of the study period (containing the temporal holdout). This is further justified in Figure 2 (right), where a shift around March 2022 leads to a higher proportion of patients discharged on Pathway 1 and a lower proportion of those discharged on Pathway 3. This highlights the need for data drift detection and frequent re-evaluation of the machine learning model if deployed in practice.

### Machine learning explanations

Explainable machine learning enables the attribution of the most important features for prediction. Since SHAP is globally consistent, the relative contribution to prediction can be stratified by target class (by grouping all predictions of a given discharge pathway). In Figure 7, we show the relative importance of admission characteristics (i.e., features) in predicting discharge needs. Features are grouped by types of information (e.g., Demographics, Patient History) and combined when encoding similar raw data (e.g., Previous Specialties is the sum of individual features ‘Last spell Specialty’ and ‘Dominant Specialty’). Supplementary Table 1 lists all individual features, and Supplementary Figure 1 shows the importance of individual features without combination.

**Figure 7:**
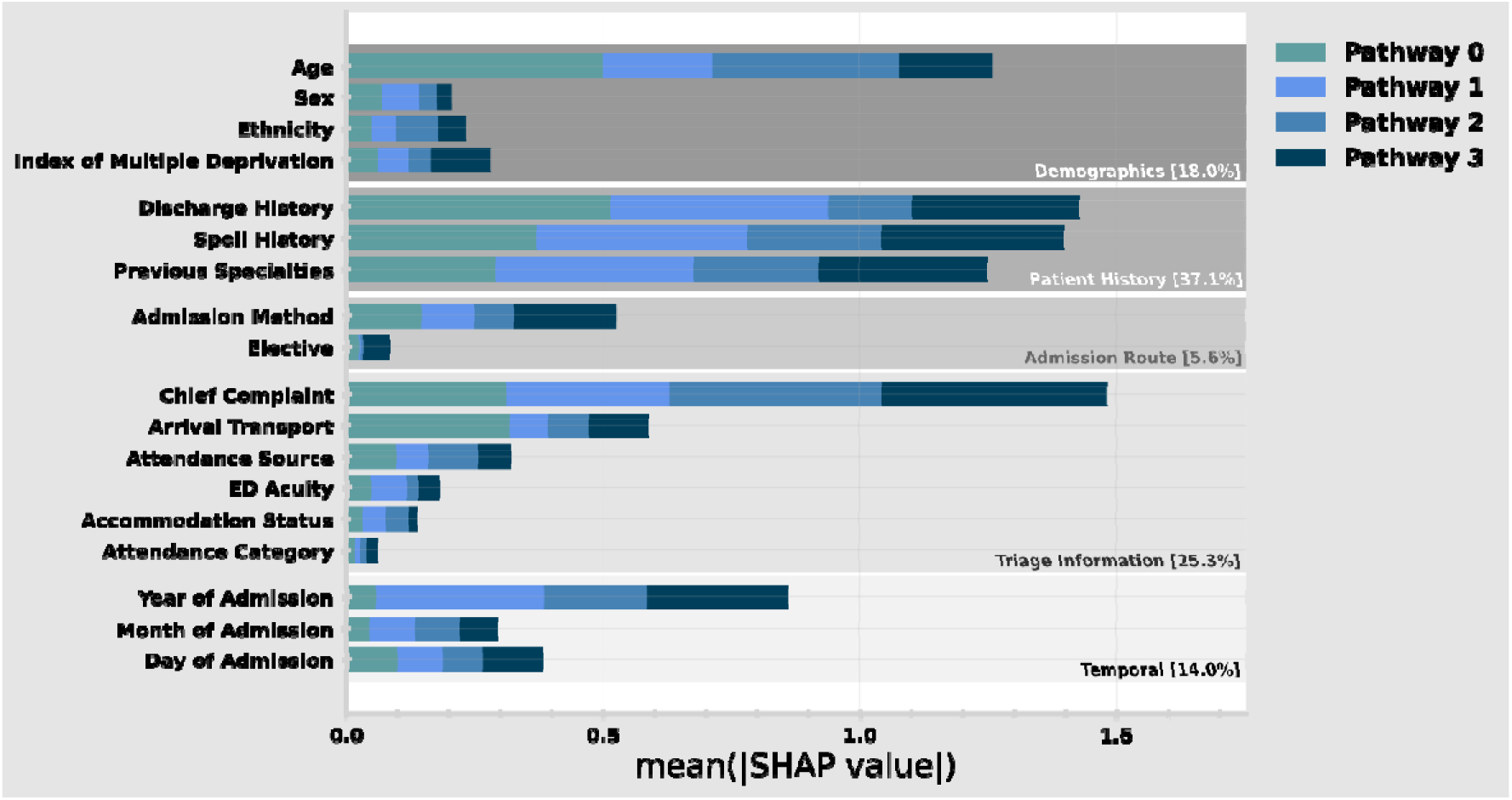
Relative importance of admission characteristics for predicting discharge pathway in the patient holdout test set. Importance is computed by normalised absolute SHAP values (i.e., larger values mean higher relative importance). The size of each complete bar shows the overall importance for prediction. Each bar is stacked so the importance for each discharge pathway can be identified (e.g., Arrival Transport is far more important for predicting pathway 0 than 1-3). Some features are grouped in categories (e.g., Discharge History contains a variety of different data encodings). An equivalent plot for the temporal holdout test set is given in Supplementary Figure 3.

We find that having a patient history (both in terms of spells and discharges) is the most significant factor in prediction, followed by the information collected at Triage in the ED. The importance of the Year of Admission reflects the importance of operational circumstances and policy at the given timeframe. Given that our study cohort contains periods still operationally disrupted by the COVID-19 pandemic, this is likely to contribute to this dependence.

Having identified the most important features for prediction, it is important to ensure our model is well generalised in the case that any of this information is not available for a particular prediction. Performing a subgroup analysis, we find our model is robust for patients without discharge history (OVR AUROC: 0.834 [0.824 0.843]), spell history (0.843 [0.827 0.859]), and for those which are not admitted via ED (0.849 [0.833 0.867]) compared with the overall model performance of 0.845 [0.838 0.852].

### Comparison to clinical predictions and hybrid model

We now compare our model to pathway predictions made by clinicians in-line with UHS standard operating procedure^13^. This corresponds to all pathway updates registered in APEX by clinicians within 24 hours of admission, usually at the first consultant review. It is important to note that only 46.7% of spells (relative to Figure 6) have clinical assessments within 24 hours of admission.

In Figure 8, we compare the performance between the machine learning model (solid), clinical updates within 24 hours of admission (dot-dashed) and the hybrid model (dashed) as described by the ROC (left) and PR (right) curves. These show the discriminative power for Pathway 0 (i.e., the ability to recognise onward discharge needs or not) through an OVR strategy (see Methods for explanation). As previously introduced, the hybrid model aims to combine the machine learning model class probabilities with the clinical prediction to derive a more powerful recommendation. We note that the AUROC and Average Precision are computed using probabilities for machine learning and hybrid models, whereas clinical pathway updates are absolute (i.e., only probability 1 for a single class), which likely understates their skill.

**Figure 8:**
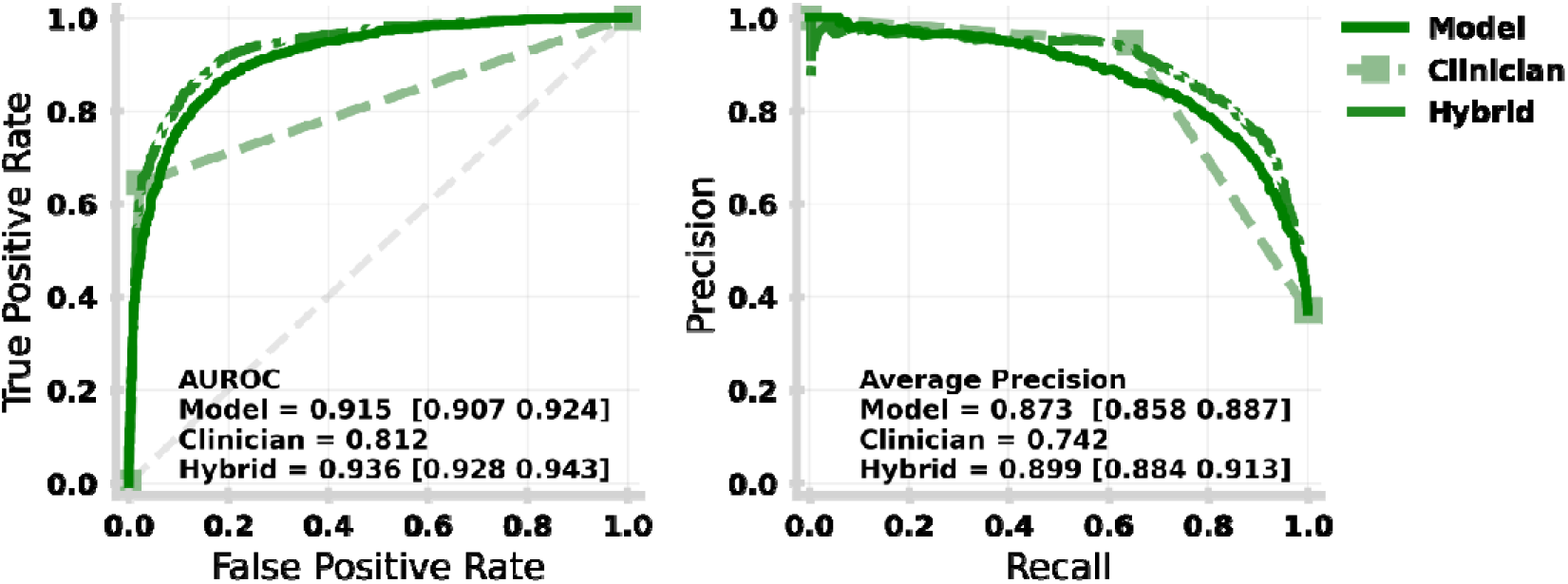
Receiving Operating Characteristic (left) and Precision-Recall (right) curves for each of the machine learning model (solid dark green), clinical pathway updates made with the first 24 hours of the spell (dashed light green) and hybrid model (dot-dashed mid green) for the patient holdout. An equivalent plot for the temporal holdout test set is given in Supplementary Figure 6.

Figure 9 shows the confusion matrices for each of the machine learning models (left), clinical predictions made within the first 24 hours of the spell (middle) and the hybrid model (right). Clinical predictions have significantly higher recall for Pathway 0, meaning clinicians are far more effective (correct 98% of the time) at identifying no onward care in comparison to the ML (86%) and hybrid (88%) models. This is a natural reflection of the tuning of the machine learning and hybrid models being weighted to improve the recall of higher acuity onward care needs coming at the cost of a higher false alarm rate. As a result, our hybrid (and machine learning only) model offers a marked improvement on identifying Pathway 2 (41% vs 15%) and Pathway 3 (49% vs 40%). Independent of tuning, we also note intrinsic differences in skill with clinicians being more effective at identifying Pathway 1, while generally struggling with Pathway 2.

**Figure 9:**
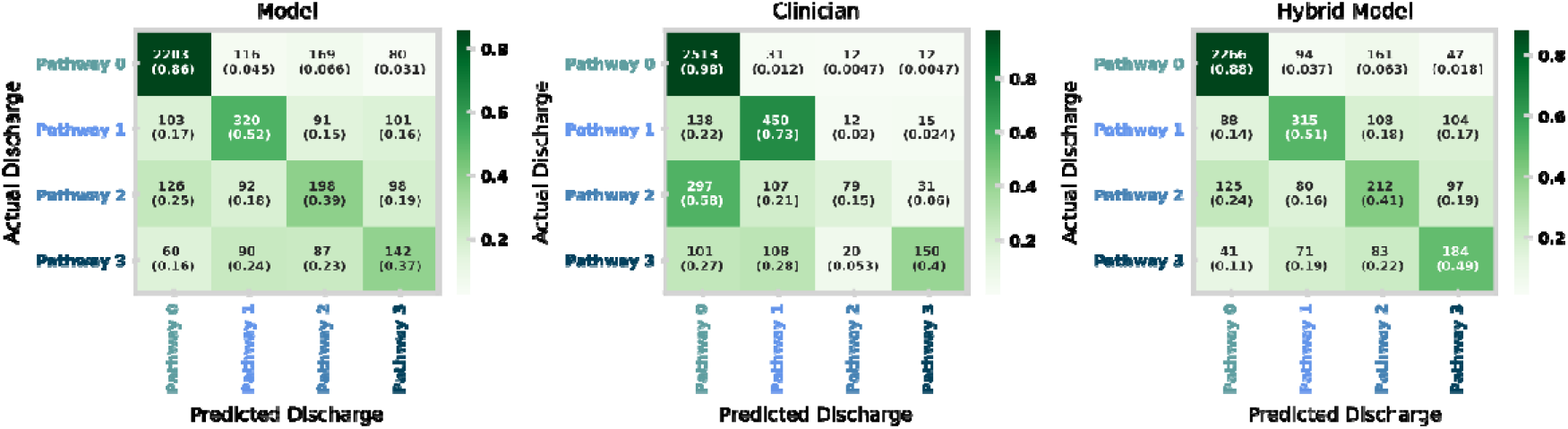
Confusion matrices for the machine learning model (left), clinical pathway updates made within first 24 hours of spell (middle) and the hybrid model (right) for the patient holdout test set. An equivalent plot for the temporal holdout test set is given in Supplementary Figure 7.

## Discussion

A significant bottleneck for patient flow through the NHS is the transfer of patients from hospital beds to social care services. Delays in discharge for medically fit patients are driven by the mismatch between bedspace pressures in hospitals and the ability of understaffed social care services to continue to accept further patients. In this work, we have demonstrated the potential for explainable machine learning to support clinical care teams with reliable information on the initial assessment of onward care needs (AUROC 0.915 [0.907-0.924], AP 0.873 [0.858-0.887] for identifying onward needs in general). We find that performance is comparable to an initial clinical assessment and can provide initial planning suggestions for spells (>50%), when clinical assessment does not occur within 24 hours of admission. Capacity pressures in hospitals compound delays by reducing the time and bandwidth of clinical teams to review and assess potential onward care requirements promptly. These predictions and subsequent model explanations could help clinical teams ensure that elevated onward care needs are identified promptly, especially as time and resource pressures prevent review.

In addition to delays due to pressures on clinical teams, pressures on social care create significant lead times in finding space for a new care patient. As a result, it is critical that onward care needs can be identified as early as possible post admission to the hospital, with the highest acuity (i.e., Pathways 2 and 3: short- and long-term intensive care respectively) most important to identify early. We identify that leveraging the clinician’s initial assessment along with the ML prediction (i.e., hybrid model) offers a significant improvement (AUROC 0.936 [0.928-0.943], AP 0.899 [0.884-0.913]) on the ability to recognise acute care needs (i.e., Pathway 2 and 3) from clinicians or ML alone (Figure 8 bottom panel).

To adapt machine learning predictions and explanations into clinical practice, the implementation of a clinical decision support (CDS) system must be considered. Clinical decision support (CDS) systems are digital tools which can support clinicians to make more equitable, evidence-based decisions^33-35^. CDS systems have been demonstrated to improve the quality of decision-making, in turn reducing errors in addition to supporting shared decision-making practice^34^. They have been applied across a variety of care and decision settings using a combination of clinical tools, technologies, information resources and guidance^33,35^. Specifically, the implementation of knowledge-based systems has been further expanded through the use of data-driven machine learning-based CDS systems^36,37^ and medical devices^38,39^. In this context, it is important to note the intrinsic differences in skill between our proposed machine learning and hybrid models in comparison to the clinical assessments. While our models are tailored to better identify high acuity Pathways (2 and 3), they have a higher error rate for Pathways 0 and 1. It is critical that care is not ‘over-prescribed’ which in turn exacerbates care placement spaces and delays arrangement of care for others.

In future work, we look to integrate our explainable machine learning model into a CDS system designed to assist complex discharge throughout a patient spell. This will involve adapting our existing supervised ML model to accept additional information as it becomes available throughout treatment (e.g., blood test results and procedures), which in turn will improve predictive performance as the spell progresses, reducing the false-alarm rate for those who only need lower acuity care. These predictions and explanations will be integrated into a clinical interface through co-design with clinical care teams and the acute discharge bureau at UHS.

### Strengths and limitations

Strengths of this research include the large and robust data collection for discharge planning, which itself was used by complex discharge teams at UHSFT; the large hospital (in terms of bed number) is NHS England. The routinely collected and digitised data is used directly for national reporting as part of the statistical snapshot for acute discharge delays aggregated by NHS England. As a result, the discharge pathway updates are accurately recorded for the cohort spells. This study also benefits from direct collaboration between hospital care teams, discharge teams and researchers to provide different perspectives.

Limitations of this research include that this study is single site (i.e., no other site validation) and performed retrospectively. There is also an absence of planning information for most hospital spells at UHSFT which are not considered in this study. Discharge planning is only created for more complex cases, and, as a result, our patient cohort is more complex than hospital averages. However, most hospital spells which were removed were short spells with 84% having a length of stay of one day or less. Consequently, these spells are unlikely to contribute to significant delay or require onward planning.

## Data Availability

Data will be made available upon reasonable request to persons with a university affiliation. Requestors will need appropriate data protection, governance, and ethical review in place.

## Acknowledgements

This report includes independent research funded by the National Institute for Health Research Applied Research Collaboration Wessex. The views expressed in this publication are those of the author(s) and not necessarily those of the National Institute for Health Research or the Department of Health and Social Care. We thank the Southampton Emerging Therapies and Technologies (SETT) Centre at UHS for support with data access and insight.

