## Supplementary Materials for "Predicting onward care needs at admission to reduce discharge delay using machine learning"

**Supplementary Material**

**S1 Supplementary Methods**

***S1.1 Data and Feature Engineering***

All numerical variables were used directly as input to the model, whereas categorical variables are target-encoded^18,19^. Target-encoding is the process of replacing a feature’s categories with the mean of the target for the given category (i.e., a supervised approach to encoding). Since there are multiple target classes, we target encode each feature for each target class. Lower cardinality categorical variables are one-hot encoded using Scikit-Learn^19^ . Ordinal variables are assigned to be integers and used directly where appropriate. The encoding of each feature is described in full below.

| **Name** | **Encoding** | **Additional Notes** |
| --- | --- | --- |
| Age at admission | N/A |  |
| Index of Multiple Deprivation Decile | N/A | Index highlighting estimation of deprivation. <https://www.gov.uk/government/statistics/english-indices-of-deprivation-2019> |
| Days since last spell | N/A |  |
| Total length of stay (4 years) |  | Sum of time spent in hospital over the prior 4 years. In Figure 7 this is grouped into ‘ |
| Total number of spells (4 years) | N/A | Total number of spells over the prior 4 years |
| Days since last discharge pathway | N/A |  |
| Last discharge pathway | N/A | Discharge pathway of most recent prior spell (max 6 months prior). 0-3 as defined by D2A. |
| Maximum discharge pathway (6 months) | N/A | Highest acuity discharge pathway in last 6 months |
| Minimum discharge pathway (6 months) | N/A | Lowest acuity discharge pathway in last 6 months |
| Total number of discharge pathways (6 months) | N/A |  |
| Admission Type | One-Hot | Whether admission was elective or non-elective |
| Sex | One-Hot | Male/Female/Unknown |
| Emergency Department Acuity | Ordinal | Emergency Severity Index (assessed acuity of patient on emergency department attendance) |
| Admission Day-of-the-week | Ordinal | i.e., Monday-Sunday encoded |
| Admission Day | Ordinal | i.e., day of month |
| Admission Month | Ordinal |  |
| Admission Year | Ordinal |  |
| Admission Method | Target | Admission route (e.g., via emergency department, GP referral, elective procedure, patient transfer). |
| Attendance Source | Target | Who referred patient (e.g., self/family, medical professional, carer). Only provided for those admitted via ED. |
| Accommodation Status | Target | e.g., home, residential care institution, homeless. Only provided for those admitted via ED. |
| Arrival Mode | Target | How patient arrived (e.g., ambulance, arranged own transport). Only provided for those admitted via ED. |
| Chief complaint | Target | Reason for attending the ED. |
| Ethnicity | Target |  |
| Last spell specialty (4 years) | Target | Primary specialty of care for patient’s most recent prior spell. |
| Dominant specialty (4 years) | Target | Most common primary specialty of care for all spells over the last 4 years. |
| Last spell discharge destination (4 years) | Target | e.g., home, residential institution, homeless. Only provided for those admitted via ED. This is not the same as last discharge pathway. |

**Supplementary Table 1:** Full list of features used in the machine learning model, how this data was encoded and additional notes.

***S1.2 Modelling***

For our machine learning model, we used the XGBoost framework, which implements a gradient-boosted decision tree approach^23^. Implementing an ensemble of weak learners, gradient boosting algorithms manage to decrease bias while preserving or lowering variance in the prediction error through iteratively creating trees fit to the residual error^24^. Gradient-boosted trees are a popular choice for large tabular data, which includes a variety of continuous, binary, and categorical data. Tree-based models can also naturally deal with missing data consistently (i.e., through the creation of splits of whether data is missing or not rather than imputation), which is important for routinely collected clinical data, as here. Here, we used a XGBClassifier with a ‘multi-softmax’ objective, other multi-class objectives were tested but resulted in consistent performance.

While gradient-boosting algorithms have been validated to be excellent at extracting signals from data, they are prone to overfitting. To help avoid overfitting, model hyperparameters were selected using five-fold cross-validation grouped at the patient level (i.e., individual patients cannot appear in different folds). On training, we also defined 50 early stopping rounds (i.e., when the train set performance metrics become better than the validation set) to prevent overfitting. A Tree-Structured Parzen Estimator Sampler was implemented with the Optuna library^25^ to find suitable hyperparameters for each of balanced accuracy, macro f1 score, and micro average precision (using 100 trials individually for each metric run). No discernible difference in performance was found between the tuning run, but hyperparameters found for Macro F1 Score were used in our models. Supplementary Table 2 describes the full results of the hyperparameter tuning.

| **Hyperparameter** | **Value** |
| --- | --- |
| n_estimators | 275 |
| max_depth | 5 |
| learning_rate | 0.167 |
| subsample | 0.538 |
| colsample_bytree | 0.377 |
| gamma | 1.66 |

**Supplementary Table 2:** Tuned hyperparameters for fiducial XGBoost model found from Tree-Structured Parzen Estimator implemented with the Optuna library.

**S2: Supplementary Results**

***S2.1 Individual feature importance***

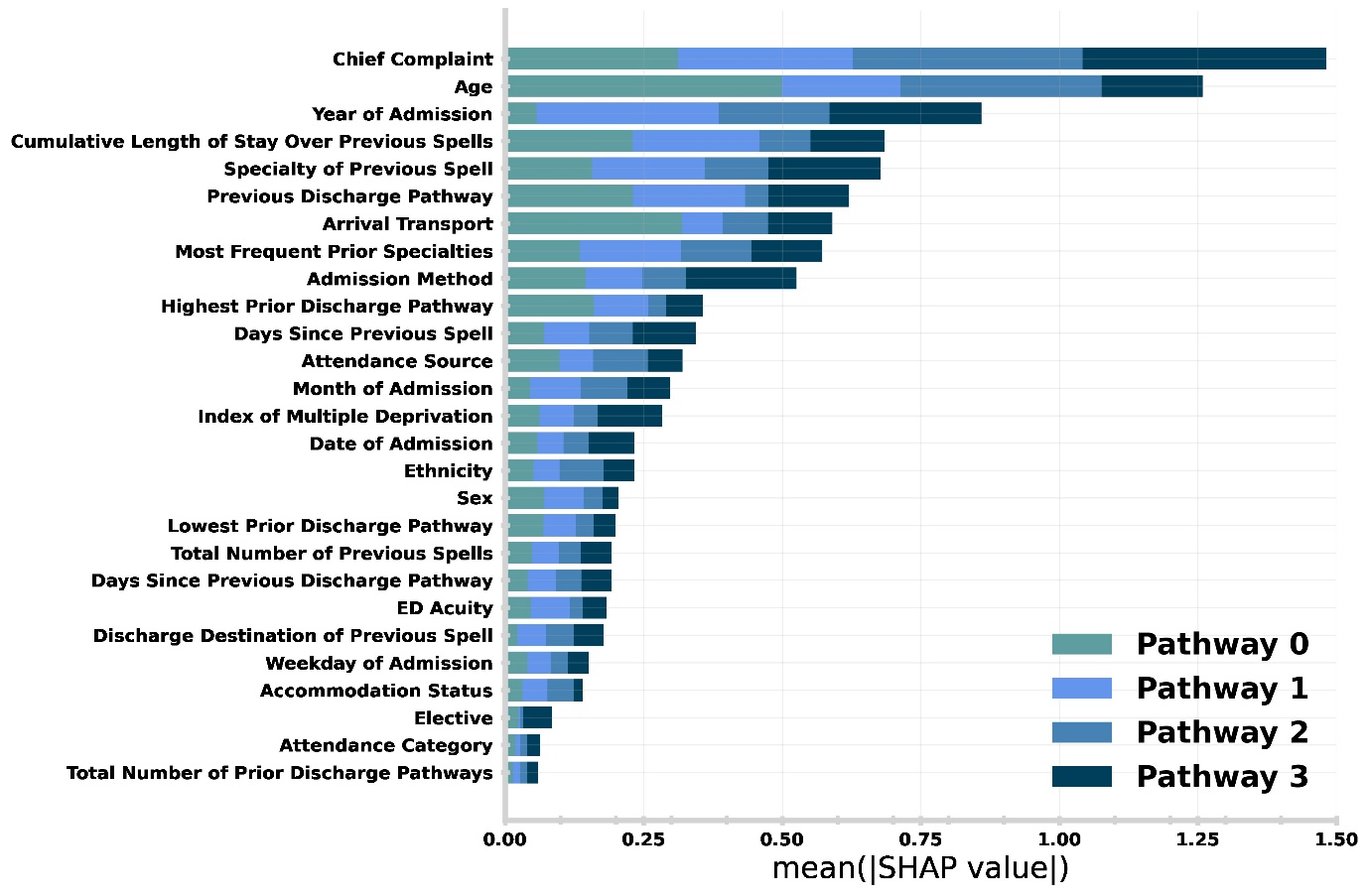

**Supplementary Figure 1:** Relative importance of admission characteristics for predicting discharge pathway in the patient holdout test set. Importance is computed by normalised absolute SHAP values (i.e., larger values mean higher relative importance). The size of each complete bar shows the overall importance for prediction. Each bar is stacked so the importance for each discharge pathway can be identified (e.g., Arrival Transport is far more important for predicting pathway 0 than 1-3).

***S2.2 Comparison to temporal holdout***

In this subsection, we present results from the temporal holdout test set (1st October 2022 to 1st January 2023). These are the conjugate plots for those presented in the main text results section. The temporal holdout is designed to emulate the real-world performance of our model.

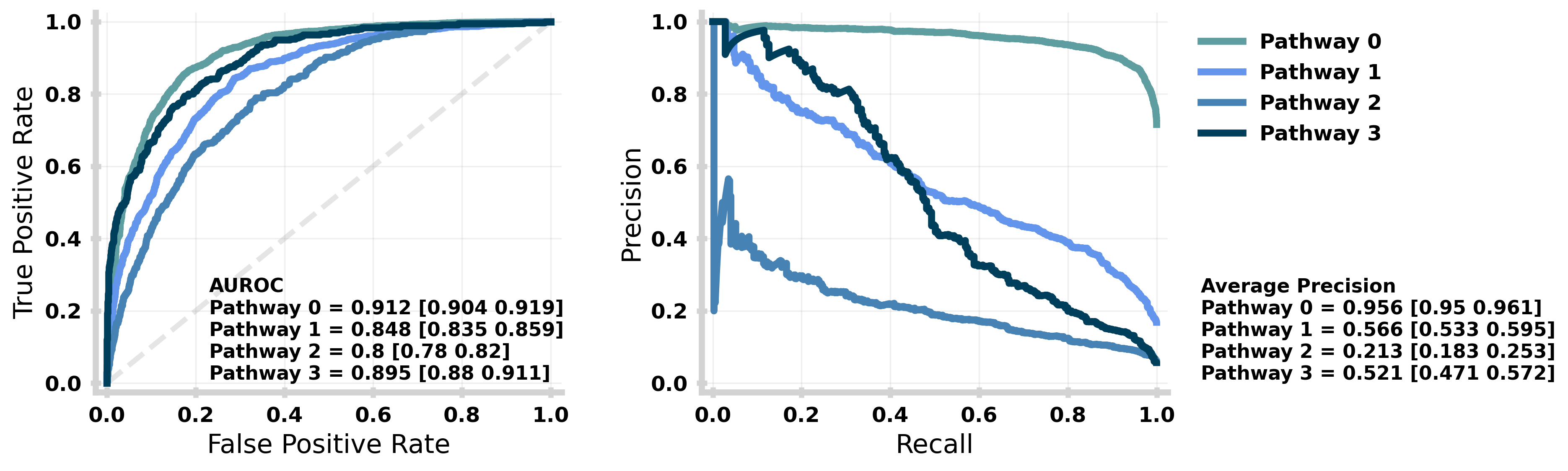
**Supplementary Figure 2**: Performance of the machine learning model stratified by target class (i.e., discharge pathway) for the temporal holdout test set. The performance for each target class is given by the receiving operator curve (left) and precision-recall curve (right) with the Area Under the ROC and Average Precision (over all recall thresholds) annotated respectively with 95% confidence intervals given in square brackets. Both ROC and PR are evaluated using a One-vs-Rest (OVR) strategy for each target class. An equivalent plot for the patient holdout test set is given in Main Text Figure 4.

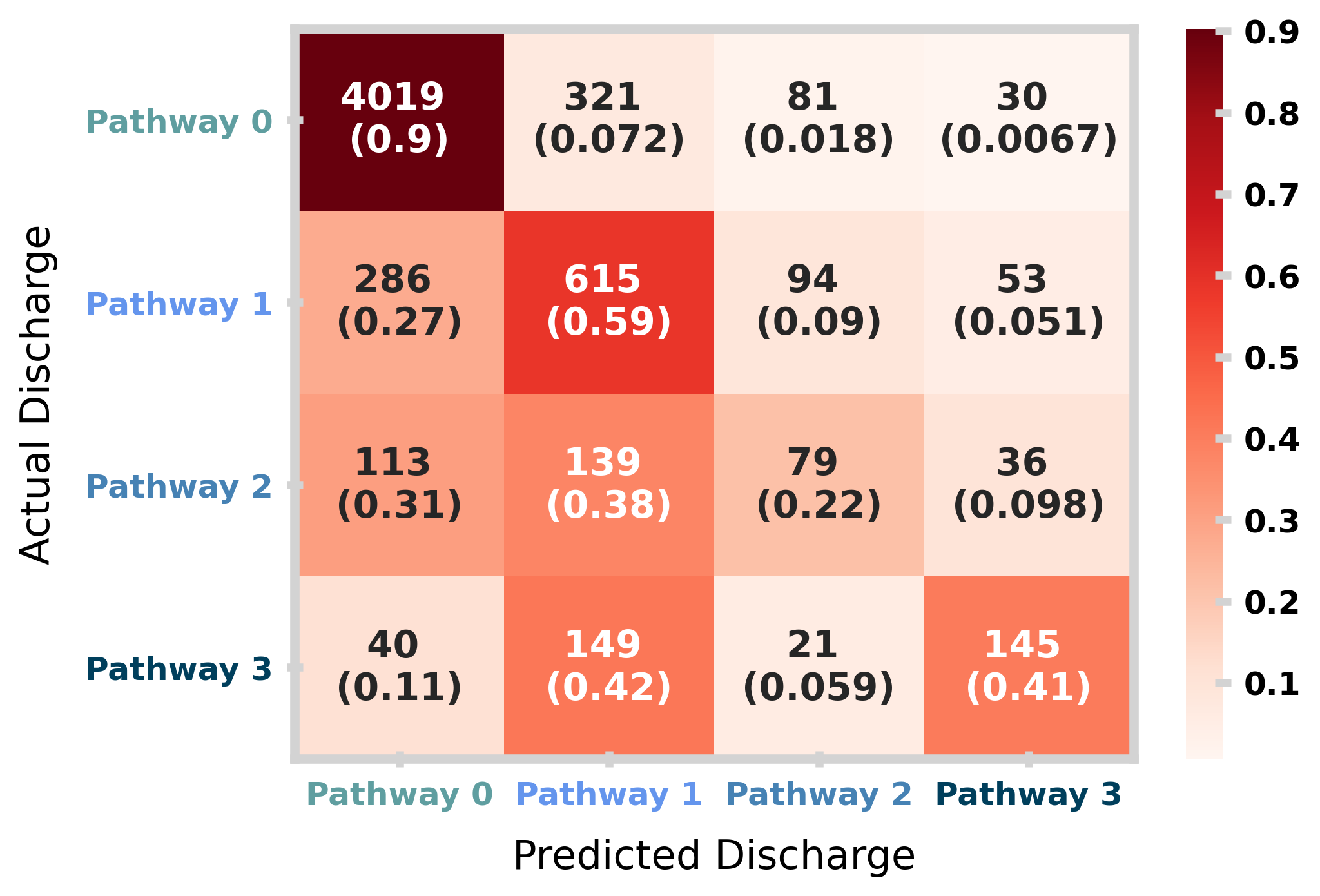

**Supplementary Figure 3**: Confusion matrix for the temporal holdout test set. The matrix shows the distribution of model prediction (columns) against the actual discharge (rows). In each square, the total counts and row normalised (i.e., by actual discharge) proportions are shown. An equivalent plot for the patient holdout test set is given in Main Text Figure 6.

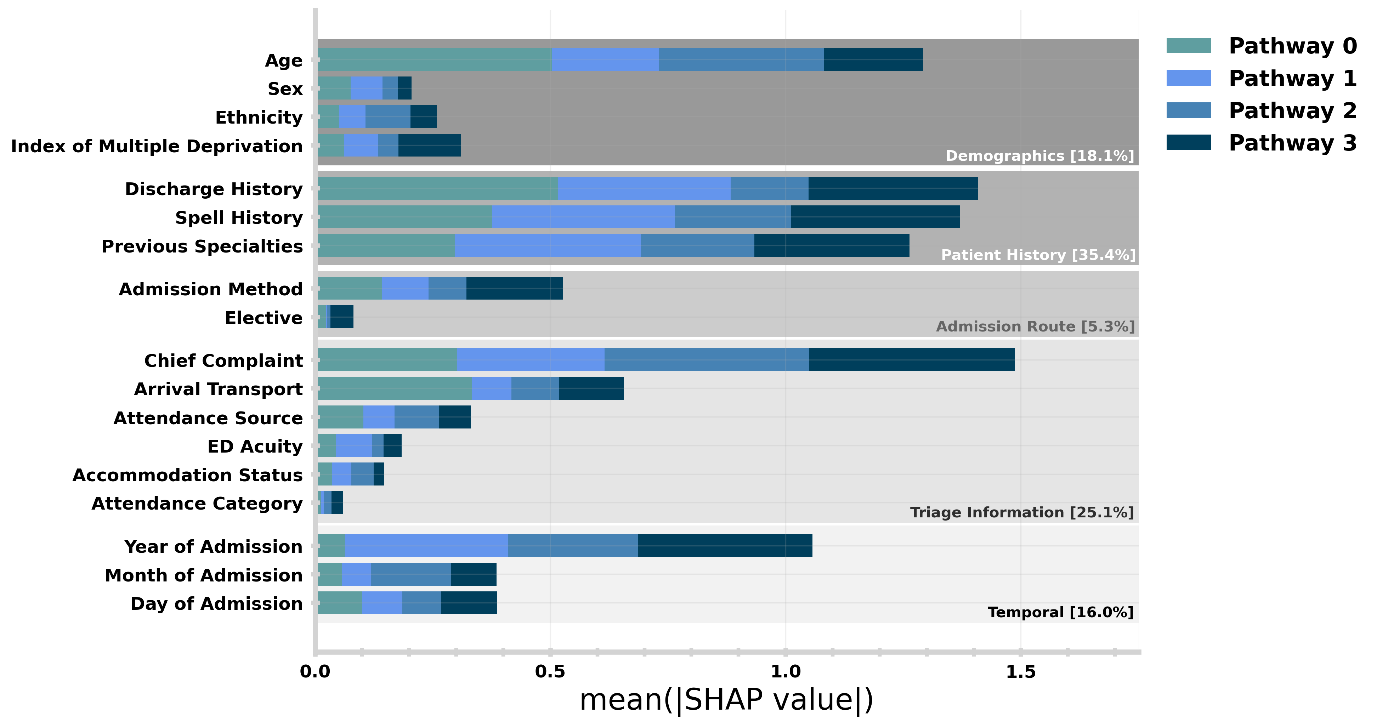

**Supplementary Figure 4:** Relative importance of admission characteristics for predicting discharge pathway in the temporal holdout test set. An equivalent plot is given for the patient holdout test set in Figure 7 in the main text.

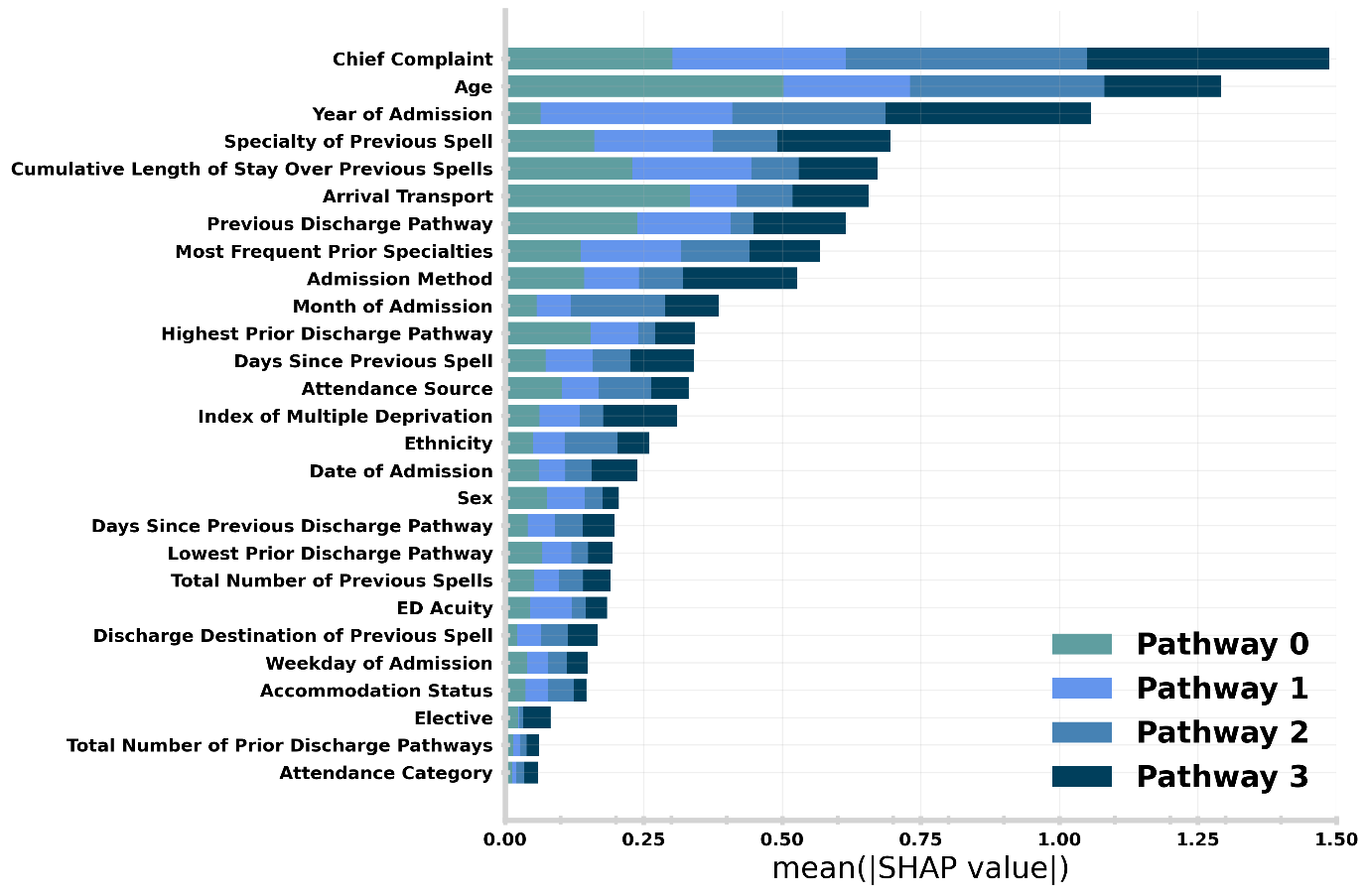
**Supplementary Figure 5:** Relative importance of admission characteristics for predicting discharge pathway in the temporal holdout test set. An equivalent plot for the patient holdout test set is given in Supplementary Figure 1.

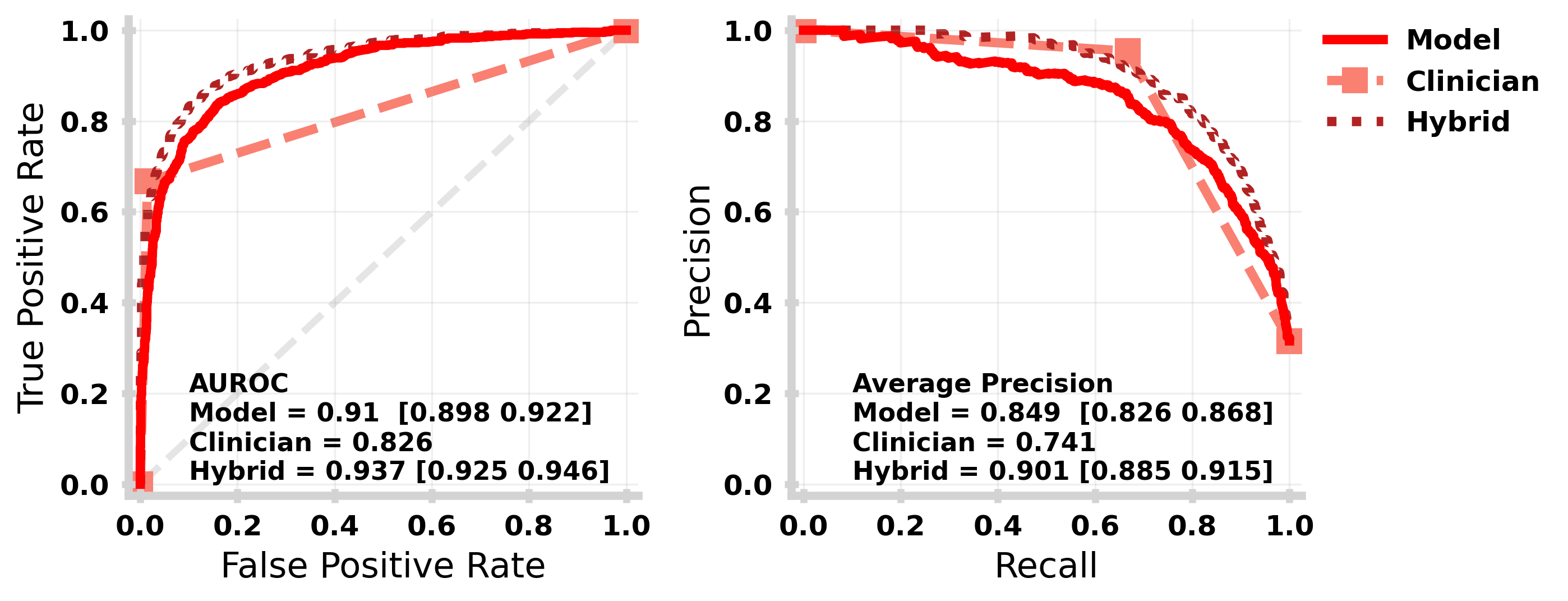
**Supplementary Figure 6:** Receiving Operating Characteristic (left) and Precision-Recall (right) curves for each of the machine learning model (solid mid red), clinical pathway updates made with the first 24 hours of the spell (dashed light red) and hybrid model (dot-dashed dark red) for the temporal holdout. An equivalent plot for the patient holdout test set is given in Main Text Figure 8.

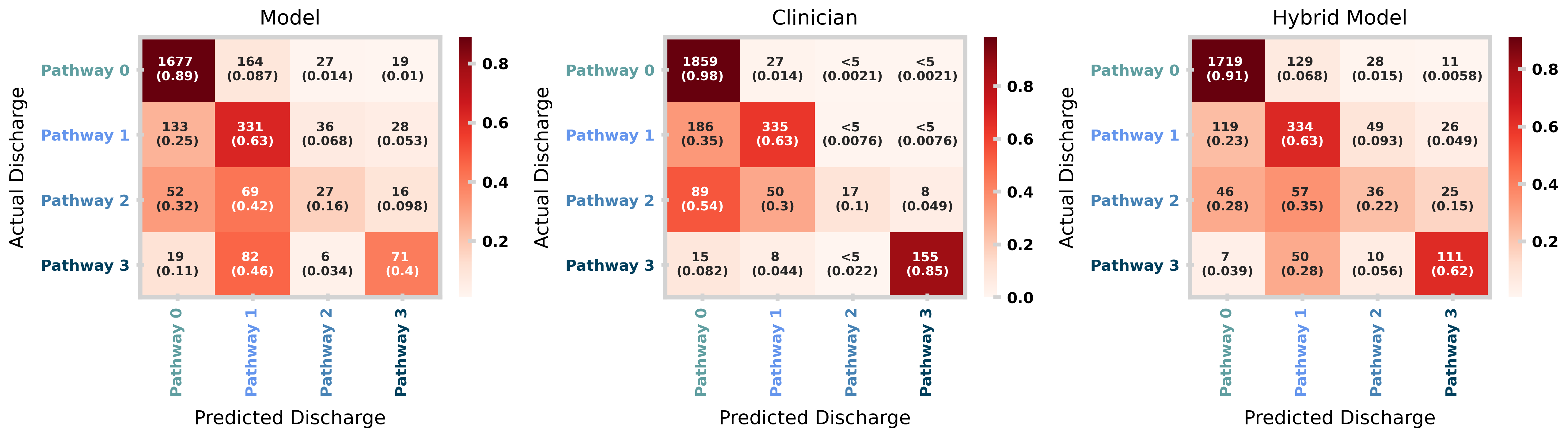
**Supplementary Figure 7**: Confusion matrices for the machine learning model (left), clinical pathway updates made within first 24 hours of spell (middle) and the hybrid model (right) for the temporal holdout test set. An equivalent plot for the patient holdout test set is given in Figure 9.
